# Chronic fatigue associated with post-COVID syndrome versus transient fatigue caused by high intensity exercise: are they comparable in terms of vascular effects?

**DOI:** 10.1101/2022.02.03.22270294

**Authors:** Michal Chudzik, Anna Cender, Robert Mordaka, Jacek Zielinski, Joanna Katarzynska, Andrzej Marcinek, Jerzy Gebicki

**Author notes:** Correspondence: Jerzy Gebicki, Institute of Applied Radiation Chemistry, Lodz University of Technology, 90-924 Lodz, Poland, or, Michal Chudzik, Medical Center, Saint Family Hospital, 90-302 Lodz, Poland.

## Abstract

**Purpose:** The pathophysiology of chronic fatigue associated with post-COVID syndrome is not well recognized. It is assumed that this condition is partly due to vascular dysfunction developed during an acute phase of infection. There is great demand for a diagnostic tool that is able to clinically assess post-COVID syndrome and monitor the rehabilitation process.

**Patients and Methods:** The Flow Mediated Skin Fluorescence (FMSF) technique appears uniquely suitable for the analysis of basal microcirculatory oscillations and reactive hyperemia induced by transient ischemia. The FMSF was used to measure vascular circulation in 45 patients with post-COVID syndrome. The results were compared with those for a group of 26 amateur runners before and after high intensity exercise, as well as for a control group of 32 healthy age-matched individuals.

**Results:** Based on the NOI and RHR parameters measured with the FMSF technique, it was found that chronic fatigue associated with post-COVID syndrome is comparable with transient fatigue caused by high-intensity exercise in terms of vascular effects. Both chronic fatigue associated with post-COVID syndrome and transient fatigue caused by high-intensity exercise are associated with vascular stress in the macrocirculation and microcirculation.

**Conclusion:** The NOI and RHR parameters measured with the FMSF technique can be used for non-invasive clinical assessment of post-COVID syndrome, as well as for monitoring the rehabilitation process.

## Introduction

Severe acute respiratory coronavirus 2 (SARS-CoV-2) is responsible for the COVID-19 pandemic, which has resulted in a global healthcare crisis. The health consequences of COVID-19 infection are broad, and not restricted to the respiratory tract. It is now generally accepted that COVID-19 infection causes serious injuries to blood vessels by damaging the vascular endothelium.^1-3^ Numerous studies have reported that patients recovering from SARS-CoV-2 infection complain of persistent symptoms, including fatigue, brain fog, diffuse myalgia, and weakness, which may lead to chronic fatigue known as post-COVID or long-COVID.^4,5^ Female sex is a risk factor associated with post-COVID syndrome, with sex hormones possibly playing an important role in that risk.^6,7^

The list of post-COVID symptoms is long, but fatigue is the most common, occurring in more than 70% of post-COVID patients. There is limited mechanistic knowledge explaining the pathophysiology of chronic fatigue associated with post-COVID. It can be hypothesized that post-COVID syndrome is primarily connected with permanent vascular dysfunction caused by initial vascular injuries occurring during the acute phase of COVID-19 infection. Therefore, it seems rational to compare the vascular effects of chronic fatigue associated with post-COVID with the vascular effects of transient fatigue caused by high intensity exercise. High-intensity exercise is known to impose transient stress on the vascular endothelium, as measured by flow-mediated dilatation (FMD).^8,9^ This effect is particularly pronounced in individuals who are not highly physically fit. It is assumed that high-intensity exercise is accompanied by a corresponding increase in reactive oxygen species (ROS), which has the potential to decrease nitric oxide (NO) bioavailability. This effect is often called the “exercise paradox”. In this sense, exercise can be considered as a stressor inducing anti-stress responses, which can be positive for physical fitness.

To test our hypothesis that the chronic fatigue associated with post-COVID is associated with permanent vascular dysfunction caused by initial vascular injuries, we decided to compare the vascular effects associated with post-COVID with those generated by high-intensity exercise. For this purpose, we applied the Flow Mediated Skin Fluorescence (FMSF) technique. The FMSF technique can be used for non-invasive assessment of vascular circulation and metabolic regulation.^10-16^ A direct comparison of the vascular effects underlying post-COVID fatigue with exercise-induced fatigue using the FMSF technique could enable the identification of diagnostic parameters for clinical assessment of post-COVID syndrome and monitoring the rehabilitation process.

## Materials and Methods

### Study Population and Clinical Characteristics

The studied population consisted of three groups: post-COVID, control, and amateur runners. The results for the post-COVID group were compared with those for the control group of age-matched healthy individuals without a history of COVID-19 infection. The vascular effects associated with transient fatigue caused by high-intensity exercise were measured in the group of amateur runners before and after exercise.

A summary of the characteristics of each group is presented below:

- **post-COVID group** 45 patients (26 female and 19 male), age range 30–50 years, after COVID-19 infection. The patients all declared having no health related problems prior to infection. The patients expressed subjective feelings of limited tolerance to exercise and above 50% greater fatigue compared to their pre-COVID-19 levels. These symptoms must have continued for at least four weeks following the last symptoms of infection. A list of the multiple symptoms associated with post-COVID syndrome with their frequencies in the group is presented in Table 1.
- **control group** 32 healthy individuals (13 female and 19 male), aged 30–50 years, without COVID-19 infection history.
- **amateur runners group** 26 highly trained amateur runners (all men), aged 21–40 years, participants of various sports competitions (long-distance running, cross-country, marathons). All athletes had valid health certificates issued by a physician specializing in sports medicine, which provided eligibility for training and competition. Exclusion criteria were illness symptoms, injuries, and taking of drugs (temporarily or chronically). The cardiopulmonary exercise test (CPET) was performed on an H/P Cosmos treadmill (H/P Cosmos Sports & Medical GmbH, Germany). The exercise protocol started with a 4-min warm-up at a treadmill speed of 6 km/h. Then, the treadmill speed was increased by 2 km/h every 3 min. The treadmill incline was 1% throughout the whole test. The test terminated if the athlete signaled his subjective exhaustion by raising one hand.

**Table 1.**
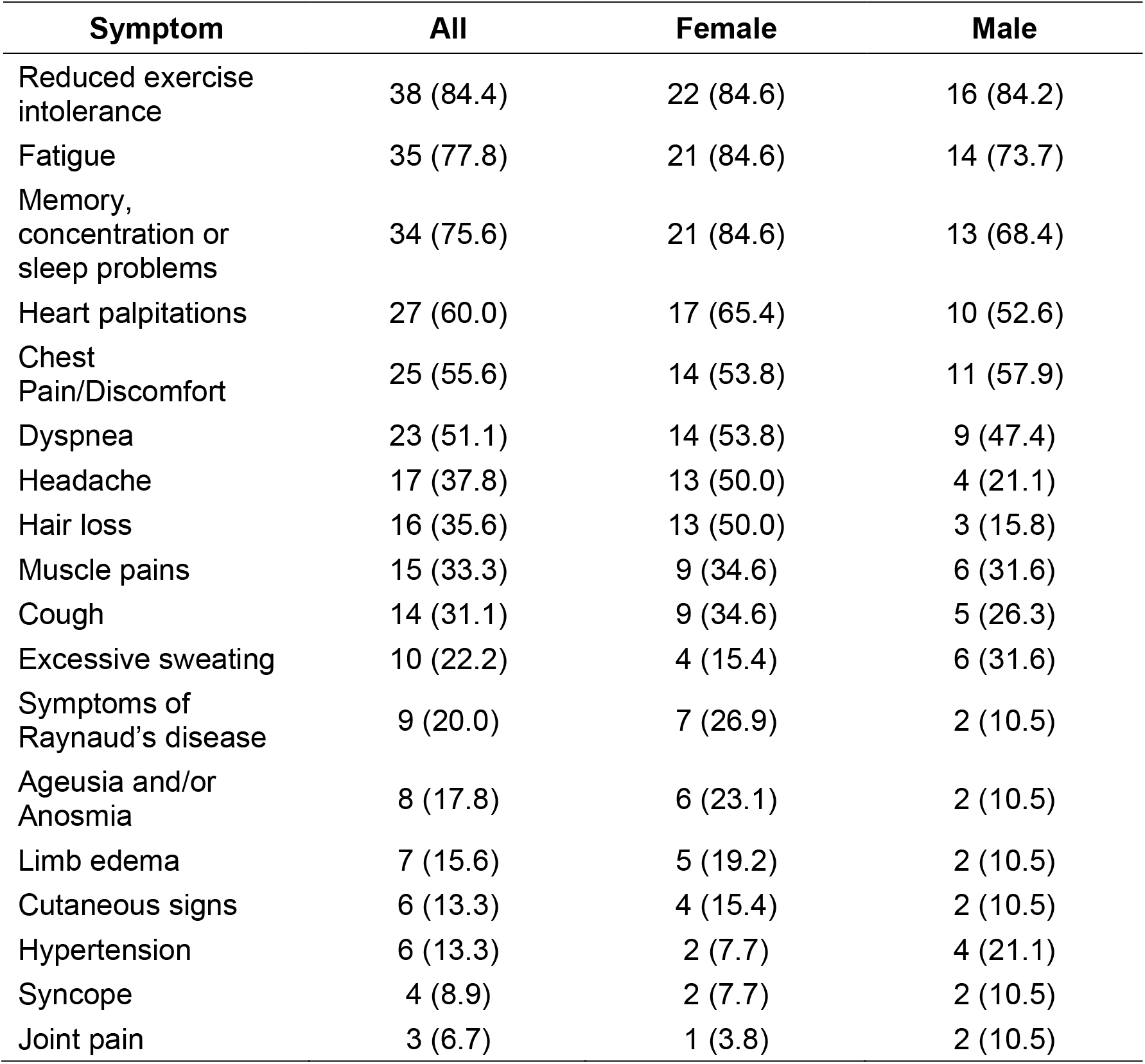
Frequencies of symptoms in individuals with post-COVID syndrome expressed as numbers and percentages (n (%))

| Symptom | All | Female | Male |
| --- | --- | --- | --- |
| Reduced exercise intolerance | 38 (84.4) | 22 (84.6) | 16 (84.2) |
| Fatigue | 35 (77.8) | 21 (84.6) | 14 (73.7) |
| Memory, concentration or sleep problems | 34 (75.6) | 21 (84.6) | 13 (68.4) |
| Heart palpitations | 27 (60.0) | 17 (65.4) | 10 (52.6) |
| Chest Pain/Discomfort | 25 (55.6) | 14 (53.8) | 11 (57.9) |
| Dyspnea | 23 (51.1) | 14 (53.8) | 9 (47.4) |
| Headache | 17 (37.8) | 13 (50.0) | 4 (21.1) |
| Hair loss | 16 (35.6) | 13 (50.0) | 3 (15.8) |
| Muscle pains | 15 (33.3) | 9 (34.6) | 6 (31.6) |
| Cough | 14 (31.1) | 9 (34.6) | 5 (26.3) |
| Excessive sweating | 10 (22.2) | 4 (15.4) | 6 (31.6) |
| Symptoms of Raynaud's disease | 9 (20.0) | 7 (26.9) | 2 (10.5) |
| Ageusia and/or Anosmia | 8 (17.8) | 6 (23.1) | 2 (10.5) |
| Limb edema | 7 (15.6) | 5 (19.2) | 2 (10.5) |
| Cutaneous signs | 6 (13.3) | 4 (15.4) | 2 (10.5) |
| Hypertension | 6 (13.3) | 2 (7.7) | 4 (21.1) |
| Syncope | 4 (8.9) | 2 (7.7) | 2 (10.5) |
| Joint pain | 3 (6.7) | 1 (3.8) | 2 (10.5) |

The clinical characteristics of the participants are displayed in Table 2.

**Table 2.** Characteristics of the studied population

| Characteristics | Control<br>(30–50 y.) | post-COVID<br>(30–50 y.) | Amateur<br>runners<br>(21–40 y.) |
| --- | --- | --- | --- |
|  | n = 32 | n = 45 | n = 26 |
| Age [years] | 37.8 ± 6.0 | 41.5 ± 5.2 | 27.7 ± 4.5 |
| Male/Female | 19/ 13 | 19/ 26 | 26/ 0 |
| Body mass index<br>(BMI) [kg/m<sup>2</sup>] | 24.9 ± 3.2 | 26.4 ± 4.8 | 23.3 ± 2.3 |
| SBP | 124.6 ± 12.6 | 131.5 ± 14.0 | 134.4 ± 7.8 |
| DBP | 79.4 ± 8.3 | 79.6 ± 9.9 | 73.3 ± 5.9 |
| Duration of post-COVID<br>symptoms [weeks] | – | 16.1 ± 9.8 | – |
| Sum of post-COVID<br>symptoms | – | 6.7 ± 3.1 | – |
| RHR [%] | 33.4 ± 6.2 | 27.6 ± 8.6 | 39.5 ± 9.7 |
| HR<sub>max</sub> [%] | 19.6 ± 3.8 | 18.6 ± 4.2 | 21.1 ± 6.1 |
| NOI [%] | 73.3 ± 19.9 | 51.6 ± 21.4 | 76.0 ± 21.3 |
| FM | 88.2 ± 81.3 | 68.5 ± 69.8 | 132.5 ± 92.1 |
| log(HS) | 1.7 ± 0.4 | 1.8 ± 0.4 | 1.8 ± 0.5 |

Continuous variables, mean ± SD; dichotomous variables, no. (%)

The study was conducted at the Medical Center, Saint Family Hospital, in Lodz (Poland), the Medical University of Lodz (Poland), and Poznan University of Physical Education (Poland). It conformed to the principles outlined in the Declaration of Helsinki and the study protocol was approved by the Bioethics Committee of Lodz Regional Medical Chamber (approval under STOP-COVID Registry), the Bioethics Committee at the Medical University of Lodz (approval no. RNN/325/17/KE and KE/09/18) and the Ethics Committee of the Poznan University of Medical Sciences (approval no. 1017/16). All the subjects gave written informed consent prior to participation.

### Brief Description of the FMSF Technique and the Measurement Protocol

Measurements were performed using AngioExpert, a device constructed by Angionica Ltd. The AngioExpert device uses the Flow Mediated Skin Fluorescence (FMSF) technique, which measures changes in the intensity of nicotinamide adenine dinucleotide (NADH) fluorescence from the skin on the forearm as a response to blocking and releasing blood flow. The skin is the largest organ of the human body, and is characterized by a specific metabolism. The epidermal layer of skin is not directly vascularized, and oxygen and nutrients are transported from the dermis by diffusion. Therefore, epidermal cell metabolism can be considered a unique and sensitive marker of early dysfunction in vascular circulation and metabolic regulation.

Details concerning the technical aspects and methodology of FMSF technique have been provided elsewhere.^10-12^ The measurement protocol in the present study was the same as applied previously.^15^ For analysis of the results obtained in the present study, two new parameters were introduced: Reactive Hyperemia Response (RHR) and Normoxia Oscillatory Index (NOI). These parameters are defined in Fig. 1b.

**Figure 1.**
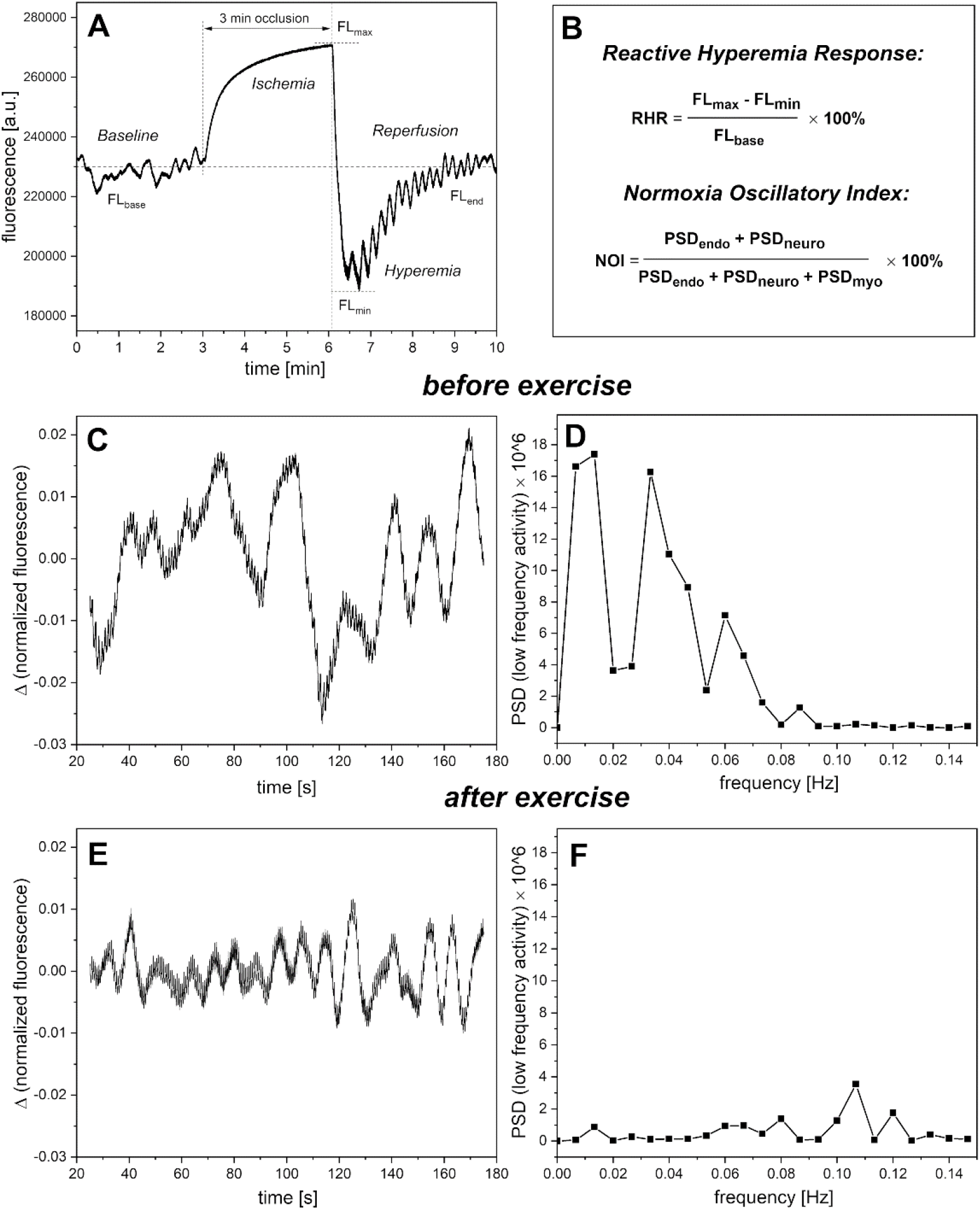
A – Exemplary FMSF trace recorded for a sportsman before high intensity exercise (male, age range 36-40 y.). B – Definition of the RHR and NOI parameters. C,D – Changes in the fluorescence signal relative to the normalized baseline before high-intensity exercise (left) and the corresponding Power Spectra Density (PSD) of the fluorescence signal in the intervals of endothelial (<0.021 Hz), neurogenic (0.021–0.052 Hz), and myogenic (0.052–0.15 Hz) activity (right); E,F – Changes in fluorescence relative to the normalized baseline after high-intensity exercise (left) and the corresponding Power Spectra Density (PSD) of the fluorescence signal in the frequency same activity intervals as for C,D (right).

The RHR parameter characterizes endothelial function related predominantly to the production of nitric oxide (NO) in the vasculature due to reactive hyperemia. RHR is a unique parameter, based on a combination of both the ischemic and hyperemic parts of the measured FMSF trace, as shown in Fig. 1a. The NOI parameter characterizes the microcirculatory oscillations detected at the baseline and represents the contribution of endothelial (<0.021 Hz) and neurogenic (0.021–0.052 Hz) oscillations relative to all oscillations detected at low frequency interval (<0.15 Hz). This parameter is particularly sensitive to high-intensity exercise. Figure 1c–d shows the changes of the fluorescence signal in the normalized baseline before high-intensity exercise, and the corresponding Power Spectra Density in the low frequency interval (<0.15 Hz). Figure 1e–f shows similar changes observed after exercise. The NOI parameter was normalized after 1–3 hours of rest and returns to the values observed before exercise. This observation is in accordance with other studies.^8,9^

We observed that both the RHR and NOI parameters should be used jointly for effective diagnostics of post-COVID fatigue and exercise-related fatigue. Based on our experience of using the FMSF technique, the NOI parameter seems more universal as it is age-independent. It is sensitive not only to fatigue caused by intense physical activity, but also to fatigue caused by emotional stress.

### Statistical Analysis

The collected data were analyzed using the dedicated FM analytical software installed on the AngioExpert device. Statistical analyses were performed with OriginPro 2018b software. The Shapiro-Wilk test was used to assess the normality of the distribution. An independent sample t-test or Mann–Whitney U test was used to compare continuous variables in the control group and post-COVID group. The paired sample t-test and the paired-sample Wilcoxon signed rank test were used to compare the FMSF parameters recorded in the amateur runners group. A two-tailed *p* < 0.05 was considered statistically significant.

## Results and Discussion

Our study indicates that post-COVID syndrome is associated with a statistically significant increase in systolic blood pressure (Supplementary, Fig. 1). A rise in blood pressure associated with COVID-19 infection has been noticed previously.^17^ Figure 2 (segments A and B) presents a comparison of the NOI parameter in the post-COVID group and the control group. A similar comparison with regard to sex differences is shown in Supplementary (Fig. 2). It is clear that post-COVID syndrome reverses the distribution of the NOI parameter, indicating that this parameter is very sensitive to the vascular problem monitored.

**Figure 2.**
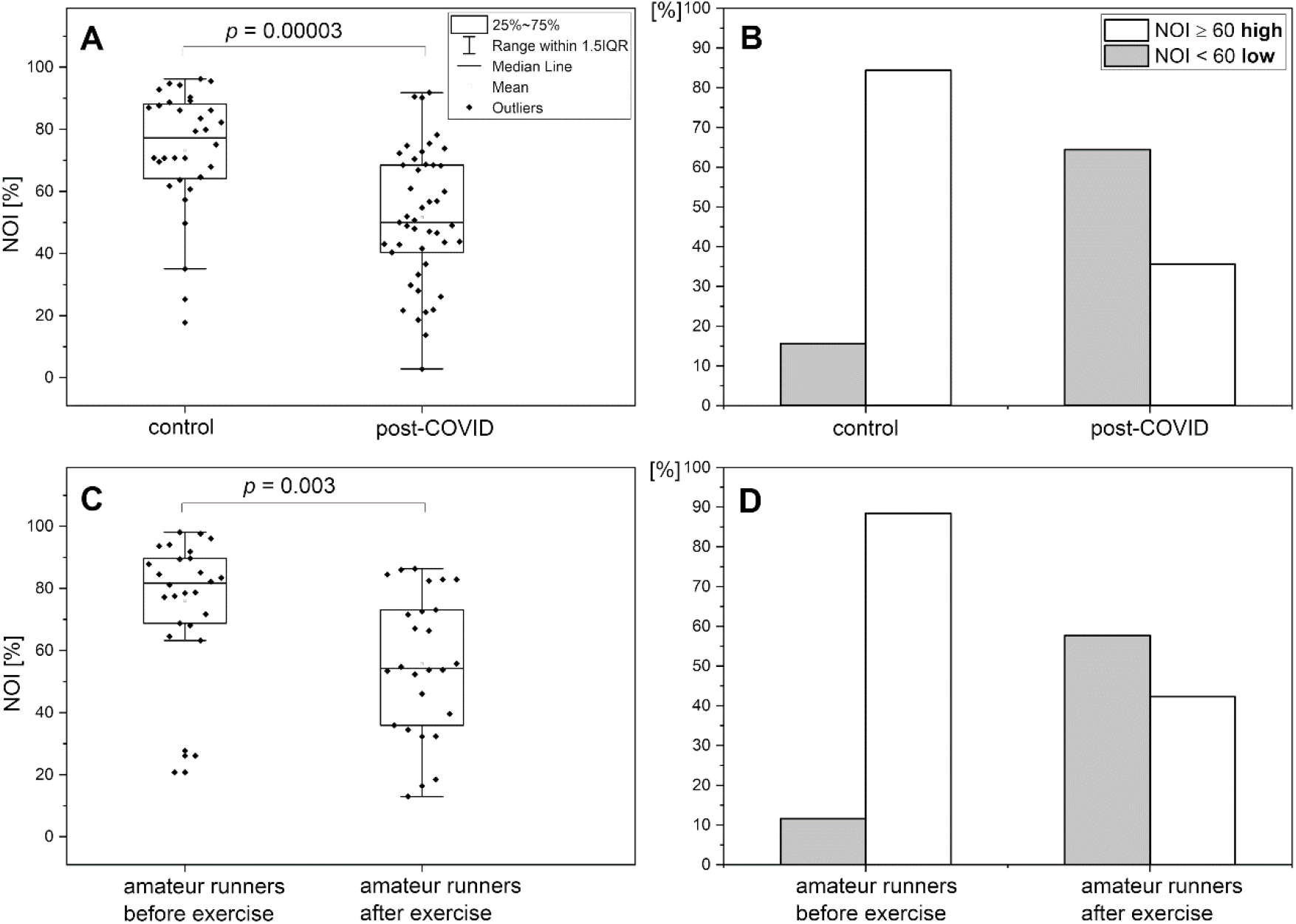
A,B – Comparison and distribution of the NOI parameter in the control group (n = 32, 19 m, 13 f, mean age 37.8 (30–50 y.)) and post-COVID group (n = 45, 19 m, 26 f, mean age 41.5 (30–50 y.)). C,D – Comparison and distribution of the NOI parameter in the group of amateur runners (n = 26, 26 m, mean age 27.7, (21–40 y.)) before and after high-intensity exercise. Differences between the parameters of the compared groups were considered statistically significant when *p* < 0.05. The *p*-values were calculated from the results of the Mann-Whitney test for comparison A and the Wilcoxon signed ranks test for comparison C.

The effects of high-intensity exercise on the NOI parameter are very similar, as shown in Fig. 2 (segments C and D). It appears that the fatigue associated with post-COVID syndrome and exercise-related fatigue exert very similar vascular effects, which can be observed as lower intensities representing endothelial and neurogenic microcirculatory oscillations at the baseline. An obvious difference is that the exercise-related effect is fully reversible after 1–3 hours of rest.

The vascular effects of post-COVID- and exercise-related fatigue measured by the RHR parameter are also quite similar, as shown in Fig. 3 (segments A and B vs. C and D) and Supplementary (Fig. 3). It should be stressed that both the NOI parameter and the RHR parameter represent distinctive properties of the vascular system. The NOI parameter characterizes microcirculation based on the measurement of microcirculatory oscillations at the baseline. In contrast, the RHR parameter characterizes the vessels based on NO bioavailability, predominantly in large and medium size arteries, due to reactive hyperemia.

**Figure 3.**
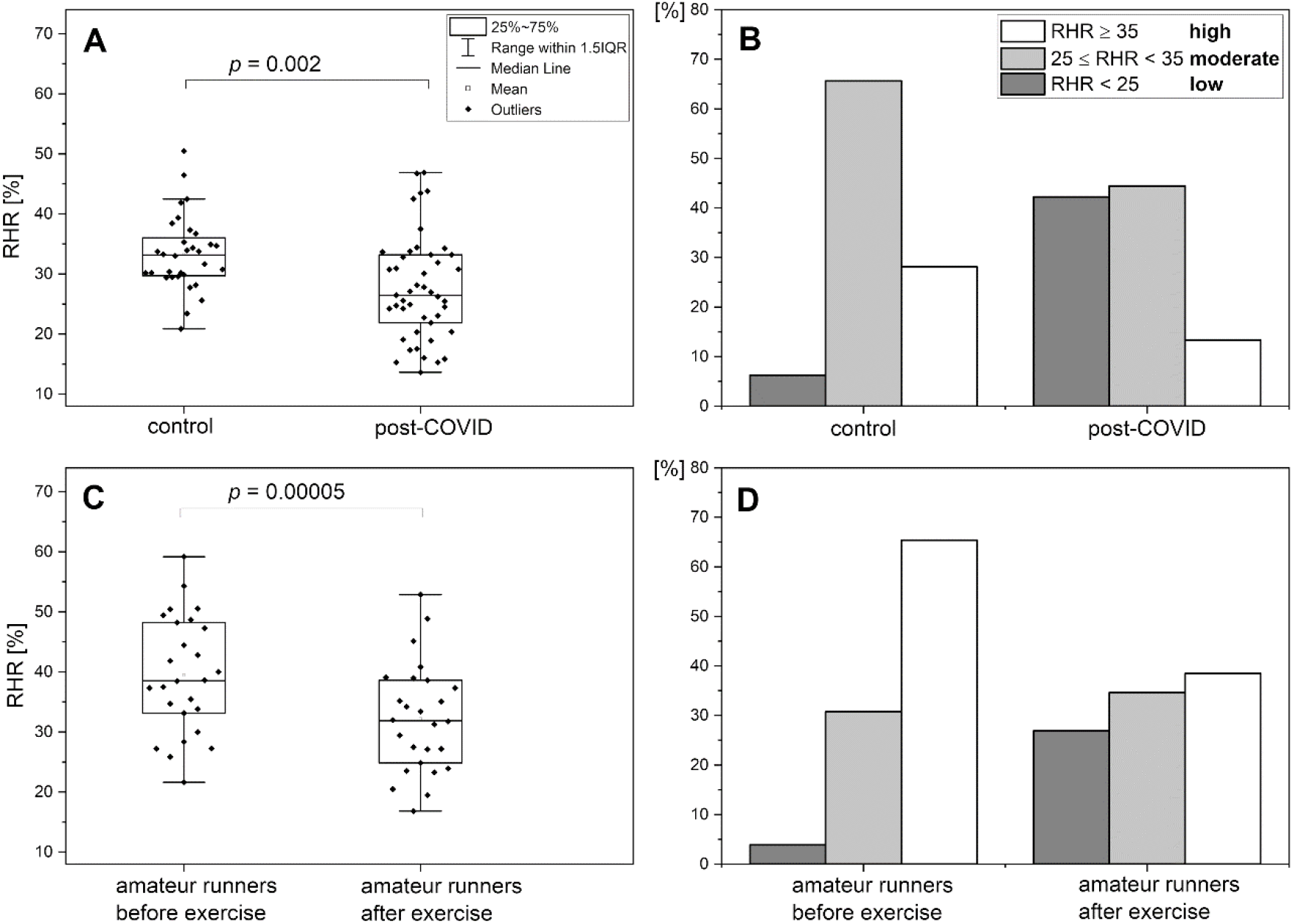
A,B – Comparison and distribution of the RHR parameter in the control group (n = 32, 19 m, 13 f, mean age 37.8 (30–50 y.)) and post-COVID group (n = 45, 19 m, 26 f, mean age 41.5 (30–50 y.)). C,D – Comparison and distribution of the RHR parameter in the group of amateur runners (n = 26, 26 m, mean age 27.7, (21–40 y.)) before and after high intensity exercise. Differences between the parameters of the compared groups were considered statistically significant when *p* < 0.05. The *p*-values were calculated from the results of a two-sample t-test for comparison A and the paired sample t-test for comparison C.

The vital conclusion is that both the macro- and microcirculation are affected by vascular stress related to both post-COVID syndrome and high-intensity exercise. Perhaps, the long list of symptoms associated post-COVID syndrome summarized in Table 1 and reported in many other studies can be linked to endothelial dysfunction present in the blood vessels of both the macro- and microcirculation.

The present study also recognizes female sex as a risk factor associated with post-COVID syndrome.^6,7^ As shown in the Supplementary Materials (Table 1), careful comparison of the RHR and NOI parameters collected for females and males seems to corroborate a difference in risk based on sex. For example, the RHR parameter is higher for females than males in the control group, but there is no such difference in the post-COVID group. Additionally, the NOI parameter seems unrelated to sex in the control group, but in the post-COVID group this parameter is lower compared to males. The apparent higher risk of females to post-COVID syndrome may indicate viral-induced sex hormone dysfunction, which can result in early menopause, as observed for other viral diseases.^6^

In previous communications, we have suggested that the Hypoxia Sensitivity (HS) parameter can be used to predict severity of the acute phase of COVID-19 infection. However, the present study indicates that this parameter has only minor utility for the diagnostics of post-COVID syndrome, as shown in Table 2.^18,19^ The explanation may be that hypoxemia plays an important role only in the acute phase of infection.

The results of this study show the utility of two key parameters (NOI and RHR) for clinical assessment and monitoring the rehabilitation process of post-COVID patients. They can also be used to identify potential treatments to improve exercise tolerance and fatigue in patients after COVID-19, as is being demonstrated by one of the co-authors.^20^

## Conclusion

The following conclusions can be drawn from this study:

- The FMSF technique appears to be uniquely suitable for the analysis of basal microcirculatory oscillations and reactive hyperemia induced by transient ischemia.
- Chronic fatigue associated with post-COVID syndrome is comparable with transient fatigue caused by high-intensity exercise, in terms of vascular effects.
- The NOI and RHR parameters measured by the FMSF technique can be used for non-invasive clinical assessment of post-COVID syndrome, as well as for monitoring the rehabilitation process.

## Data Availability

The details concerning the presented results can be obtained from the correspondence authors

