## Supplementary Material for "Chronic fatigue associated with post-COVID syndrome versus transient fatigue caused by high intensity exercise: are they comparable in terms of vascular effects?"

**Table 1.** Characteristics of the control and post-COVID groups by sex.

| Characteristics | Control (30-50 y.) | | Post-COVID (30-50 y.) | |
|---|---|---|---|---|
| | Female n = 13 | Male n = 19 | Female n = 26 | Male n = 19 |
| Age [years] | 39.0 ± 6.6 | 36.9 ± 5.5 | 41.2 ± 4.5 | 42.0 ± 6.0 |
| Body mass index (BMI) [kg/m$^2$] | 23.8 ± 3.0 | 25.6 ± 3.3 | 25.1 ± 5.4 | 28.3 ± 3.2 |
| SBP | 116.6 ± 12.3 | 130.0 ± 9.8 | 127.4 ± 14.3 | 137.0 ± 11.9 |
| DBP | 76.1 ± 7.9 | 81.7 ± 8.0 | 77.1 ± 10.0 | 83.1 ± 9.1 |
| Cigarette smoking | 1 (7.7) | 1 (5.2) | 2 (7.7) | 4 (21.1) |
| Duration of post-COVID symptoms [weeks] | – | – | 15.7 ± 8.1 | 16.5 ± 11.8 |
| Sum of post-COVID symptoms | – | – | 7.2 ± 3.2 | 6.0 ± 2.8 |
| RHR [%] | 35.5 ± 6.6 | 31.9 ± 5.6 | 27.9 ± 7.6 | 27.3 ± 10.0 |
| $HR_{max}$ [%] | 21.1 ± 4.4 | 18.5 ± 3.0 | 18.2 ± 4.3 | 19.0 ± 4.3 |
| NOI [%] | 72.8 ± 23.8 | 73.6 ± 17.4 | 47.8 ± 20.5 | 56.7 ± 22.1 |
| FM | 141.4 ± 104.1 | 51.8 ± 26.8 | 85.2 ± 83.1 | 45.7 ± 36.6 |
| log(HS) | 1.9 ± 0.4 | 1.6 ± 0.4 | 1.8 ± 0.3 | 1.6 ± 0.4 |

Continuous variables, mean ± SD; dichotomous variables, no. (%)

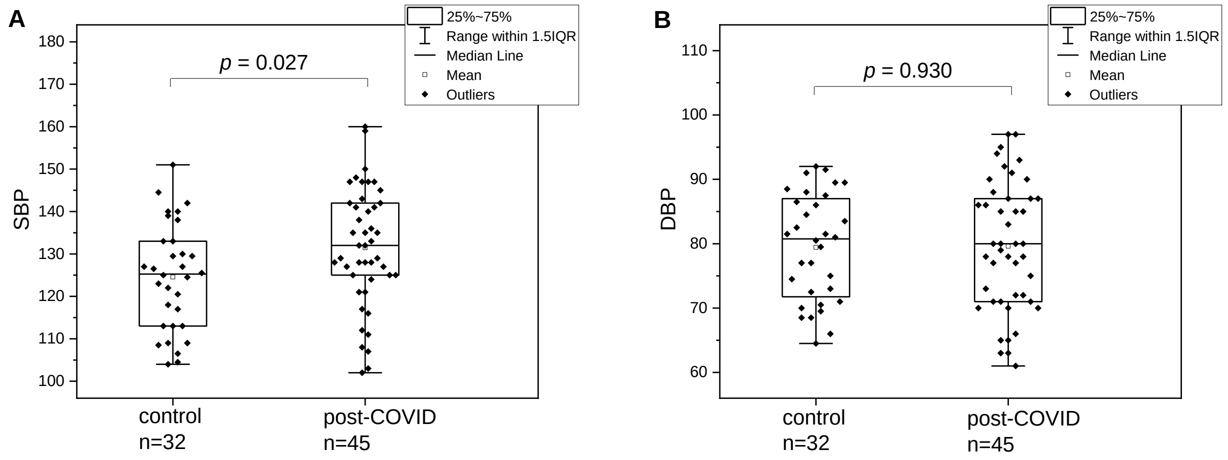


**Figure 1**. Comparison of systolic (A) and diastolic (B) blood pressure for the control group and post-COVID group. Statistical significance was set at $p < 0.05$. The *p*-values were calculated from the results of a two-sample t-test.

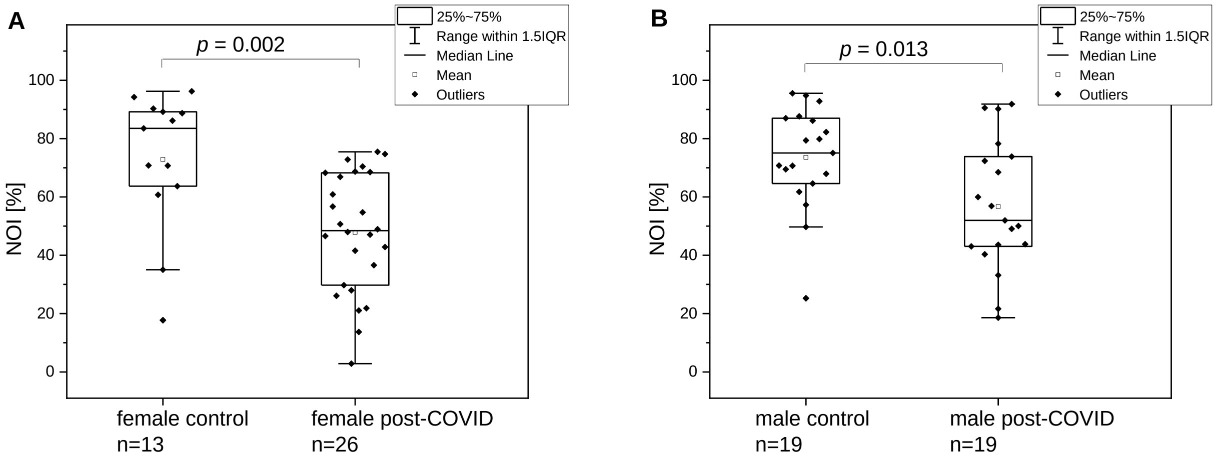


**Figure 2.** Comparison of the NOI parameter for the control group and post-COVID group by sex (A – female, B – male). Statistical significance was set at $p < 0.05$. The *p*-values were calculated from the results of a Mann-Whitney test for comparison A and the two-sample t-test for comparison B.

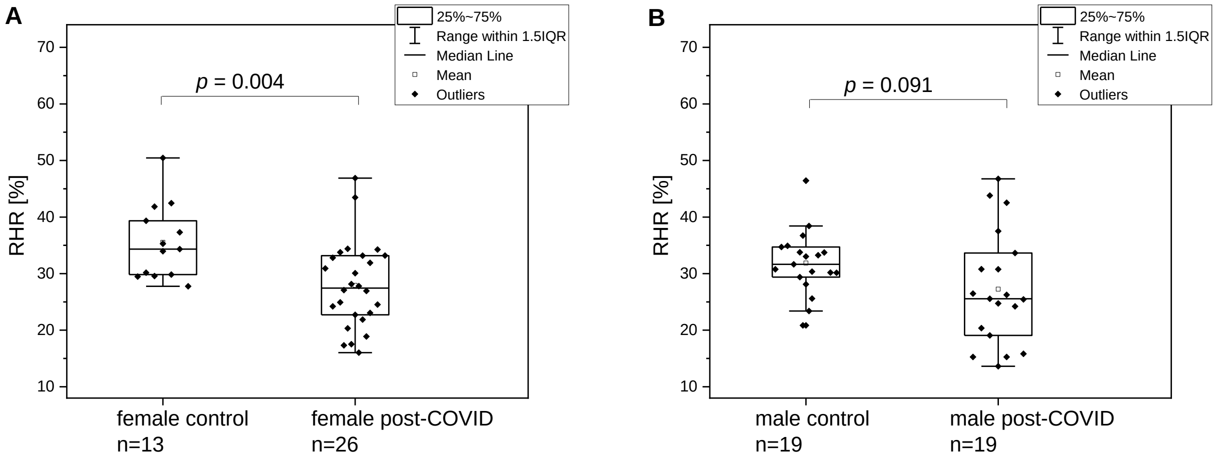


**Figure 3.** Comparison of the RHR parameter for the control group and post-COVID group by sex (A – female, B – male). Statistical significance was set at $p < 0.05$. The *p*-values were calculated from the results of a two-sample t-test.
